# Artificial intelligence-enabled analysis of UK and US public attitudes on Facebook and Twitter towards COVID-19 vaccinations

**DOI:** 10.1101/2020.12.08.20246231

**Authors:** Amir Hussain, Ahsen Tahir, Zain Hussain, Zakariya Sheikh, Mandar Gogate, Kia Dashtipour, Azhar Ali, Aziz Sheikh

## Abstract

**Background:** Global efforts towards the development and deployment of a vaccine for SARS-CoV-2 are rapidly advancing. We developed and applied an artificial-intelligence (AI)-based approach to analyse social-media public sentiment in the UK and the US towards COVID-19 vaccinations, to understand public attitude and identify topics of concern.

**Methods:** Over 300,000 social-media posts related to COVID-19 vaccinations were extracted, including 23,571 Facebook-posts from the UK and 144,864 from the US, along with 40,268 tweets from the UK and 98,385 from the US respectively, from 1^st^ March - 22^nd^ November 2020. We used natural language processing and deep learning based techniques to predict average sentiments, sentiment trends and topics of discussion. These were analysed longitudinally and geo-spatially, and a manual reading of randomly selected posts around points of interest helped identify underlying themes and validated insights from the analysis.

**Results:** We found overall averaged positive, negative and neutral sentiment in the UK to be 58%, 22% and 17%, compared to 56%, 24% and 18% in the US, respectively. Public optimism over vaccine development, effectiveness and trials as well as concerns over safety, economic viability and corporation control were identified. We compared our findings to national surveys in both countries and found them to correlate broadly.

**Conclusions:** AI-enabled social-media analysis should be considered for adoption by institutions and governments, alongside surveys and other conventional methods of assessing public attitude. This could enable real-time assessment, at scale, of public confidence and trust in COVID-19 vaccinations, help address concerns of vaccine-sceptics and develop more effective policies and communication strategies to maximise uptake.

## Introduction

The imminent availability of COVID-19 vaccines poses a pressing need to continually monitor and better understand public sentiments in order to develop baseline levels of confidence in vaccines and enable identification of early warning signals of losses in confidence [1]. This will help address the concerns of vaccine sceptics [2,3,4] and develop required public trust in immunisation [5,6] to realise the goal of herd immunity [7].

Traditionally, governments use surveys to understand public attitude; however, these typically suffer from small sample sizes, closed questions and limited spatio-temporal granularity. In order to overcome these limitations, we argue social-media data can be used to increase numbers and enable real-time analyses of public sentiments and attitudes with considerable spatiotemporal granularity. Over half the world population, including around 70% of both the UK and US populations are active social-media users, with significantly increased usage reported during the pandemic, e.g. by 37% for Facebook [8]. Since social-media data is largely unstructured, it is amenable to application of established AI techniques, such as machine learning (ML), deep learning (DL) [9] and natural language processing (NLP) [10], to extract topics and sentiments from the social-media posts.

Sentiment analysis involves categorising subjective opinions from text, audio and/or video [10] to obtain polarities (e.g. positive, negative, neutral), emotions (e.g. angry, sad, happy) or states of mind (e.g. interested versus uninterested) towards target topics, themes or ‘aspects’ of interest [11]. A complementary approach, termed stance detection [12], assigns a stance-label (favourable, against, none) to a post towards a specific predetermined target, which in itself may not be referred to, or be the target of opinion in the post. Such approaches are currently under-utilised in healthcare. In particular, there is significant untapped potential in drawing on AI-enabled social-media analysis to inform public policy research.

## Methods

### Ethics

We conducted a thorough assessment of the privacy risk to individuals posed by our research, in light of [13,14], to ensure compliance with relevant sections of the General Data Protection Regulation (GDPR). Further we have striven to comply with best practices for user protection [15,16], ensuring no non-public material is included in our dataset.

### Data Sources

We used data from both Facebook and Twitter, two of the most popular and representative social-media platforms [8][17]. We used English-language Facebook posts and tweets that were posted in the UK and the US from 1st March to 22nd November 2020. Facebook posts were obtained through the CrowdTangle platform [18], and Twitter posts from a publicly available Twitter API. We utilised hydrated tweets from the global COVID-19 dataset available at [19], which collects up to 4.4 million tweets per day including retweets, and up to 1.1 million cleaned tweets without the re-tweets. The total number of tweets hydrated and utilised for this study were over 158 million. The Facebook posts and tweets were *thematically* filtered for both COVID-19 and vaccine related keywords and then geographically filtered for the UK and the US. The first step filtering with COVID-19 related keywords utilised widely used terms from [19] (see Appendix Section-A1). The vaccination terms used for second step filtering were selected by our team: vaccine, vaccination, immunise, immunize, immunisation and immunization. The two-step thematic filtering process was applied using these keywords before processing and analysis.

### Analysis

The filtered dataset was initially pre-processed (e.g. removing links, hashtags, stop words) and a new hierarchical hybrid-ensemble based AI model was developed for thematic sentiment analysis. This utilised an average-weighting ensemble [20] of two lexicon-based methods: Valence-Aware Dictionary and sEntiment-Reasoner (VADER) [21] and TextBlob [22],. These were combined with a pre-trained DL-based model: Bidirectional Encoder-Representations from Transformers (BERT) [23], using a rule-based ensemble method, as illustrated in Figure 1.

**Figure 1:**
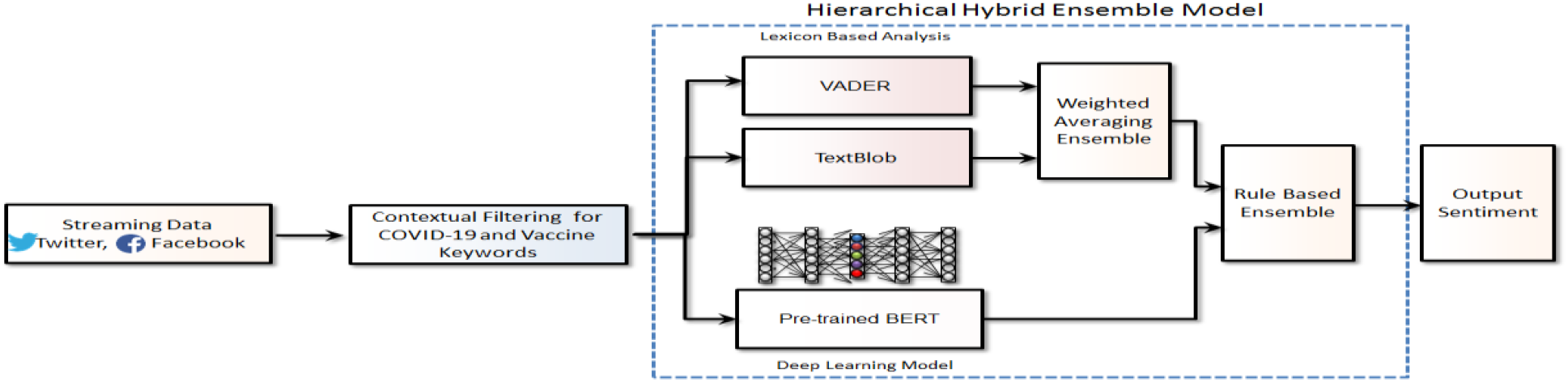
Hierarchical hybrid-ensemble based AI model and data pipeline for thematic sentiment analysis

A random 10% sample of posts and tweets were then manually annotated by the team, and checked against our hybrid-ensemble AI model’s sentiment classifications for refinement and validation. The hybrid-ensemble model was optimised based on the validation results, with sensitivity and specificity analysis showing that the lexicon-based methods provided generally better accuracy for positive sentiments, and the BERT model generally provided better performance for neutral and negative sentiments. The final rule based hybrid-ensemble method utilised the classification output of the BERT model for neutral and negative sentiments, while the weighted-average output of lexicon-based methods was selected for classifying positive sentiments.

A number of established NLP techniques were used to analyse the processed data (see details in Appendix Section-A2). Specifically, in addition to analysing averaged sentiment trends and their geo-spatial mappings in the UK and US, we statistically analysed the trends (using Pearson’s r) and compared findings with independent surveys. Sentiment word cloud and N-gram analysis was applied to specific time-periods of interest, around points of inflexion on sentiment trend graphs, to identify topics of discussion and glean insight into the positive and negative content of online discourses. The analysis was also carried out over the full period of study, to identify underlying themes and topics. Findings were validated, and further insights obtained, through a manual reading by our team, of randomly selected posts around target points of interest. Relevant social-media datasets and outputs were anonymised, and statistical aggregates made openly available for transparency and reproducibility (including through a publicly available dashboard [24]).

## Results

### Temporal Sentiment Trends

Monthly volume trends of the filtered UK and US Facebook and Twitter posts used for the target period of study are shown in Appendix (Figure-A1). Figure 2 illustrates the averaged (weekly) positive, negative and neutral Facebook sentiments from March-November 2020, for the UK and US. We identified topics of discussion around points of interest on the graphs. These are referred to in our descriptive analysis of the graphs below, and some are highlighted in Figures 2 and 3. It was interesting to note that on Facebook, the difference between the averaged positive and negative sentiment trends was more pronounced, compared to Twitter.

**Figure 2:**
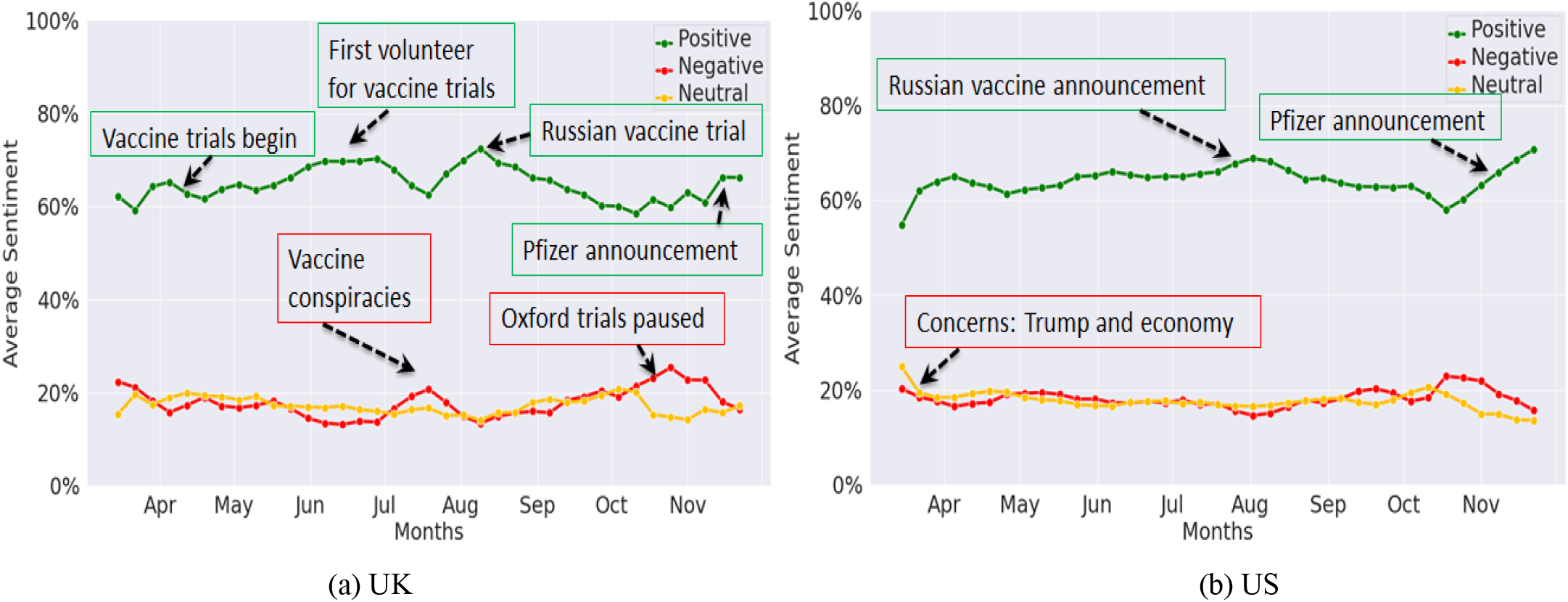
Averaged weekly Facebook sentiment trends for (a) UK and (b) US

**Figure 3:**
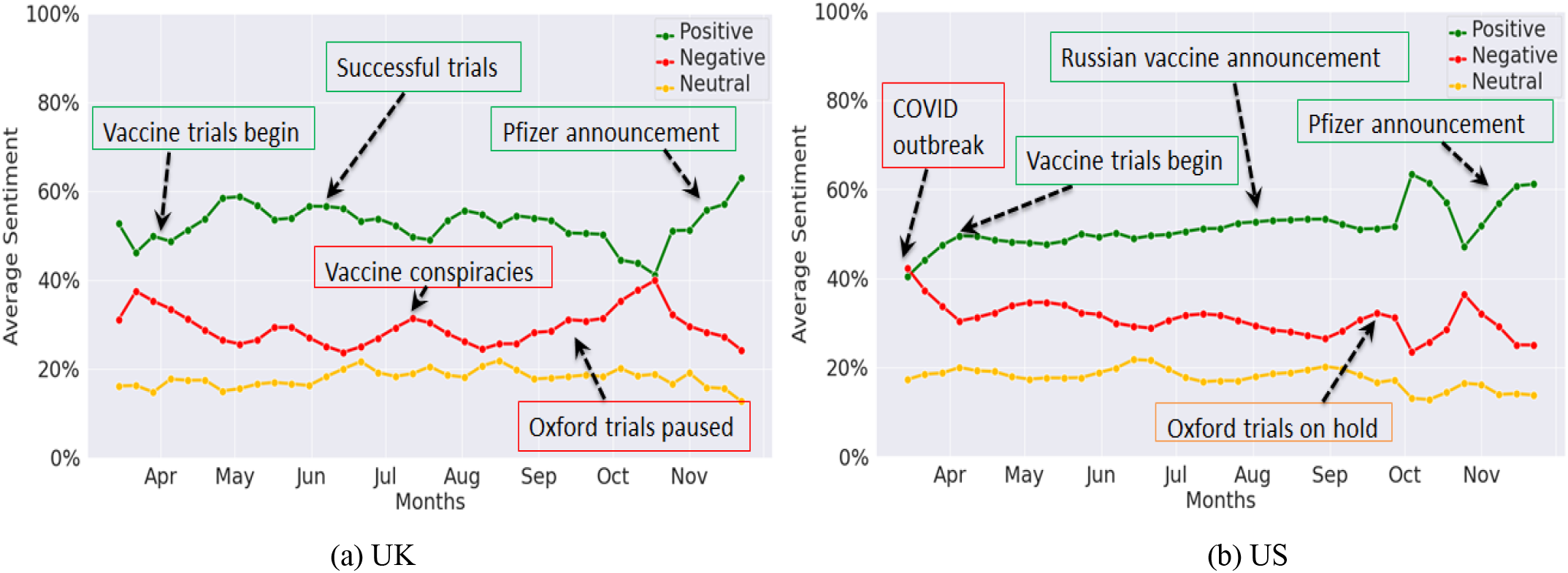
Average weekly Twitter sentiment trends for (a) UK and (b) US

For the UK, the positive Facebook sentiment trend graph was most prevalent (Figure 2(a)), and rose steadily since May, corresponding to the initiation of vaccine trials and the recruitment of the first volunteer. There was a peak in mid-August, possibly associated with news on vaccine developments in the UK and Russia. The negative sentiment trend graph inversely followed the positive trend, and discourse was centered around vaccine conspiracies and halting of trials. For the US, the positive Facebook sentiment trend was again most prevalent, and had a small peak in August, associated with posts relating to research on COVID-19 vaccine. Moreover, the negative sentiment trend graph had slightly increased in mid-September, associated with posts relating to the COVID-19 vaccine being rushed (Figure 2(b)). More recently, from mid-October onwards, there has been a growing positive sentiment trend for both the UK and the US in part attributed to the announcements from Pfizer and Moderna of successful vaccine trials [25].

Figure 3 illustrates the positive, negative and neutral Twitter sentiments from March to November 2020, for the UK and the US. We can see that in the UK (Figure 3(a)), positive Twitter sentiment was the most prevalent and its trend graph had small peaks at the end of April and July 2020. The former was found to be related to the first human vaccination trial. The negative sentiment trend had peaks in July and October 2020, simultaneous to the UK opting out of the European Union vaccination scheme, and the Oxford vaccination trials being paused [26] due to safety concerns respectively. In the US (Figure 3(b)), positive sentiment was also more prevalent, and had a major peak from end September to end November. This was related to claims by President Trump on a vaccine being ready in a few weeks, and an increase in twitter discourse due to his reference to the “herd mentality”. A small peak in the negative trend graph in mid-September was related to pausing of the Oxford vaccination trials.

For both the UK and the US, there has been a noticeable marked increase in the positive sentiment trend, since end-October, which we found related to recent breakthrough announcements by Pfizer and Moderna. Analysis of social media conversation indicated public optimism, with trial results being hailed as “good” and “amazing”, there being “hope” for the “new year” (see Appendix Figures A2 and A3). A notable peak in the negative sentiment trend graphs for both countries, around mid-October, was associated with the growing anti-vaccination movement, and with concerns around “fake news” and “misinformation”.

### Statistical Analysis of Sentiment Trends

Statistical findings are detailed in the Appendix Section-A3, which assessed the strength of the association between the predicted sentiment in the trend graph and the accuracy of the labelled data. In conclusion, there was a stronger sentiment on Twitter in relation to COVID-19 vaccinations for the US, with both positive and negative sentiments demonstrating stronger increasing and decreasing trends, as compared to the UK. Public sentiment on Facebook was found to represent decreasing positive sentiment and increasing neutral sentiment for both the UK and the US, with positive sentiment demonstrating a slightly stronger decreasing trend in the UK than the US.

### Sentiment Word Clouds and Text N-Gram Analysis

We applied these techniques to the entire period of study, to identify and analyse notable events that were of interest to social-media users, and summarise them in the Appendix: Section-A4, Figures A2, A3 and Tables A1, A2 (some of these were also identified in the analysis above, on the sentiment trend graphs).

### Geo-spatial Sentiment Analysis

The geo-spatial mapping of overall (averaged) sentiments to states in the US are shown in Figure 4 (left) and indicate that most states have a negative sentiment. The states with an overall negative sentiment towards COVID-19 vaccination were found to be concentrated in the West and Midwest of the country, namely: Idaho, Kansas, New Hampshire, West Virginia, Alabama. The states with an overall positive sentiment were in the East, namely, Maine, Colorado, Georgia, Hawaii

**Figure 4:**
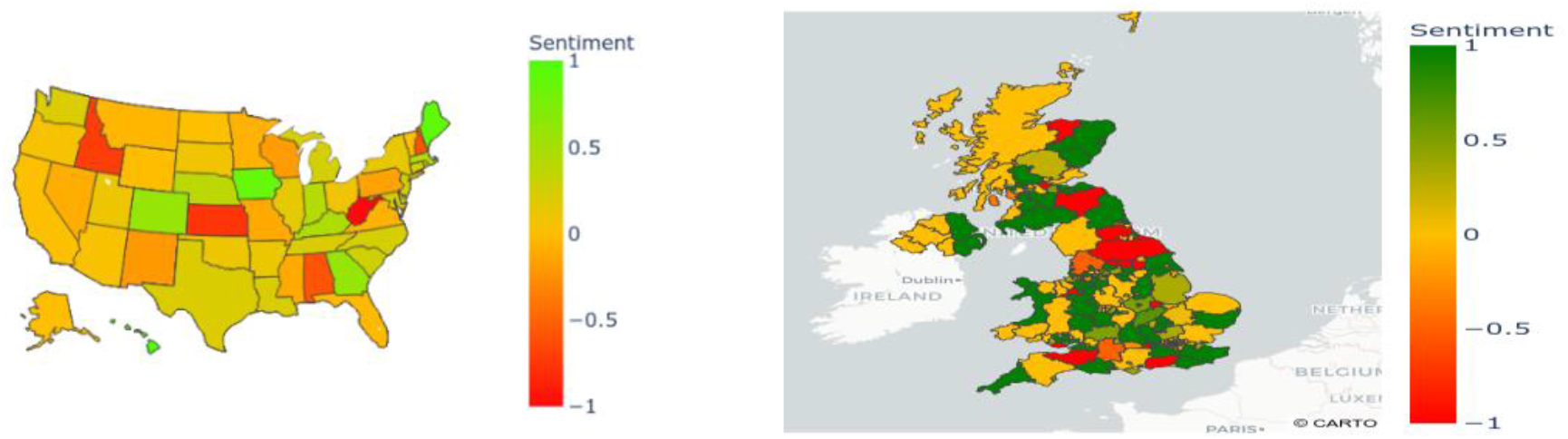
Geo-spatial mapping of averaged social-media public sentiment in US (left) vs UK (right) related to COVID-19 vaccination (+1 or green indicates positive sentiment, 0 neutral, and −1 or red indicates negative sentiment)

The geo-spatial mapping of averaged sentiments to counties in the UK is illustrated in Figure 4 (right). In contrast with the US, most counties in the UK had an overall positive sentiment towards COVID-19 vaccination. The counties with the most positive sentiment included Cornwall, Kent, East Sussex, Surrey and Dorset all in England, and Aberdeenshire, Angus and Stirlingshire all in Scotland. Furthermore, the counties with the most negative sentiment were West Sussex, Somerset, North Yorkshire and Durham, all in England.

### Overall Averaged Sentiment

Overall averaged sentiments in the UK and US on Facebook and Twitter are shown in Figure 5 below (and described in Appendix Section-A5).

**Figure 5:**
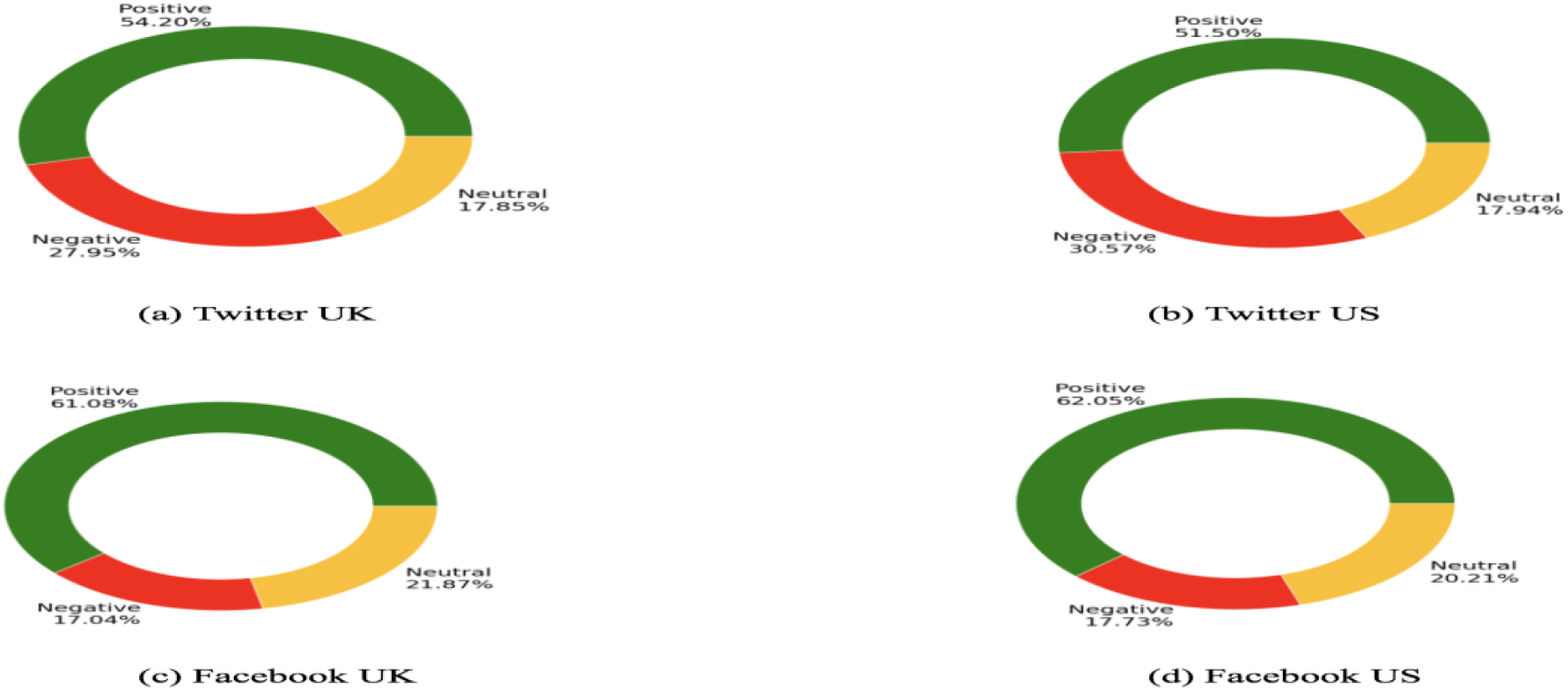
Overall averaged sentiment: (a) Twitter UK (b) Twitter US (c) Facebook UK (d) Facebook US

## Discussion

We analysed temporal variations in public sentiments on COVID-19 vaccination in the UK and the US. Key events impacting positive, negative and neutral sentiments were identified, evaluated and mapped to the temporal trends. We also mapped spatial variations in public sentiment to regions and states of the UK and the US respectively. Our geo-spatial mappings can help identify areas with more negative sentiment towards a COVID-19 vaccine which can be further studied for potential interventions, to allay underlying public fears and concerns.

Our analysis has shown that online public discourse on Facebook and Twitter platforms across both the UK and the US is evolving, with both complementary and contrasting insights gleaned from the two popular platforms. Comparative analysis results indicate that over the nine-month period of study, averaged public sentiment towards COVID-19 vaccines has been mostly positive and similar in both the UK and the US across both platforms (57.70% average across both platforms for UK vs 56.80% for the US). Positive sentiment was found to be related to public opinion on vaccine development, related trials and news related to the availability of a vaccine.

The overall averaged negative sentiments, on both platforms, were also found to be similar for the UK (22.50%) and US (24.10%). It is interesting to note that Twitter sentiment appears to be more negatively biased, with the proportion of negative sentiment being almost double that of Facebook, for both the UK (27.95% vs 17.04%) and also the US (30.57% vs 17.73%), potentially reflecting their respective user demographics. This also appears to be consistent with the study in [17], which found public opinion on Twitter to be often more negatively biased than on Facebook, where it is more positively biased. Negative sentiments in our study were found to relate to public apprehensions and concerns around delays or pauses in vaccine trials, vaccine safety, corporations and governments influencing vaccine availability and rights exclusivity for economic benefits.

A comparative analysis with independent surveys was carried out. Our findings relating to averaged positive and negative sentiment trends across both the UK and US were found to correlate broadly. In the US, during the early stages of the pandemic, polling indicated that a significant minority had low trust in a vaccine – for example, a Yahoo News/YouGov survey in May [27] found only 55% of Americans planned to get vaccinated against COVID-19, whilst almost one in five (19%) would not get vaccinated. A similar survey in July [28] indicated 42% would get vaccinated (27% would not get vaccinated), whilst a survey in September [29] found only 36% were certain they would get vaccinated (32% would not get vaccinated). More recently, an Axios-Ipsos survey in November reported, consistent with our social-media analysis findings, a marked increase in those likely to get vaccinated (51% were ‘very’ or ‘somewhat’ likely to get the first generation vaccine, increasing to 70% if the vaccine was proven to be safe and effective by public health officials) [30].

In the UK, a YouGov survey in June [31] found 41% of respondents would ‘probably’ or ‘definitely’ get vaccinated, whilst one in six (16%) of respondents ‘definitely” or ‘probably’ would not get vaccinated. The survey also found individuals who used social-media more than traditional media as their source for news, were nine percentage points less likely to be in favour of being vaccinated. A more recent UK YouGov survey in November [32], in relation to the Pfizer COVID-19 vaccine, found 67% were ‘very’ or ‘fairly’ likely to take the vaccine when available, with one in five (21%) unlikely to take it. Whilst there has been a slight increase in the proportion unlikely to take the vaccine, the proportion of those likely to get vaccinated has increased, indicating a decrease in the number of those who were previously unsure. This could be attributed in part to the recent announcements by vaccine manufacturers, and our results corroborate this with a marked increase reported in positive sentiment since mid-October, in both the UK and US, across the two social-media platforms. Further study into the change in sentiment could help us better understand factors that have contributed to this, in particular the impact of government education programmes.

It is important to consider the limitations of our data sources and techniques, and related challenges and opportunities they present for required future research. Whilst we attempted to gauge country-wide public sentiments in the UK and US, by analysing English-language posts on both Facebook and Twitter, our data may not be representative of the broader population. Users are known to differ in their social-media platform preference and usage, based on their socio-demographics (e.g. age, socio-economic status, political affiliation). Vaccinations are likely to be preferentially targeted at older populations and possibly ethnic minorities, communities which have historically lower rates of vaccination uptake (e.g. [33,34]). Further exploration is therefore imperative to increase our understanding of public perception towards vaccines, and their underlying behavioral determinants [35]. Social network analysis [36] can be used in conjunction with DL methods to effectively identify sources of fake news/misinformation and their social networks, to help deal with ‘infodemic’ challenges [37]. Demographic information, such as age, gender, race and geographic origin can also be inferred from user social-media profiles using AI techniques [38]. This can help categorise distinct groups and inform the development of demographic-level engagement and tailored communications strategies to promote diversity and inclusion in vaccination campaigns. These can also effectively account for the fact that there are genuine knowledge voids being filled by misinformation [35].

Technical limitations of our approach include challenges in determining the geographical location of users, and issues relating to the accuracy of the AI techniques (e.g. dealing with sarcasm, implicit context). The two-step keyword-based thematic filtering process and use of geo-tagged posts in the study resulted in relatively small sample sizes. This could be improved by using more sensitive filtering and data-driven search mechanisms, network meta-data (such as likes, retweets) and additional social-media and web platforms. On account of the current limitations, our approach should only be used in conjunction with other techniques for understanding public sentiments, such as focus groups, input from civil society organisations, surveys and public consultations.

Future studies could consider conducting periodic public surveys over the period of interest being explored by social-media analysis. This would ensure both methodologies were informed by each other over the course of the study to enable more fine-grained spatio-temporal analysis, allowing more robust comparisons from reciprocal findings and deeper insights for policy makers. These could also complement other qualitative methods, such as in-depth interviews and ethnographic studies as part of mixed-study approaches. Manual annotation/labelling of datasets is imperative when training AI models for NLP tasks, to ensure accuracy and generalisability. These can be affected by the skill of annotators and the proportion of the dataset that is labelled. Confounding factors, such as political affiliations, should also be included in future studies, by applying further filters to search strategies and through targeted demographic analysis, to better understand underlying determinants of public sentiment. Attitudes towards different vaccine manufacturers could also be explored, to identify and assess effective public engagement strategies to build support for ethical principles, and maximise uptake of the imminently available vaccines.

One of the main threats to the resilience of vaccination programmes globally is the rapid and global spread of misinformation. The public’s confidence in COVID-19 vaccine is known to be exacerbated by unproven vaccine safety scares seeding doubt and distrust. There are also cases where vaccine debates have been purposefully polarised, exploiting the doubting public and system weaknesses for political purposes, while waning vaccine confidence in other places may be influenced by a general distrust in the government and scientific elites. Recent surveys and polls in the UK and the US have indicated the fragility of support for vaccination, which furthers the case for a better understanding of underlying public concerns and attitudes, both at scale and in real time. Retrospective analysis of two popular and most representative social-media platforms in this study demonstrates the potential of AI-enabled real-time social-media monitoring of public sentiments and attitudes to help detect and prevent such fears, and also to enable policymakers better understand the reasons behind why some social groups may be reluctant to be vaccinated against COVID-19. This can inform more effective policy making and promote participatory dialogues around complex vaccine deployment issues, under conditions of uncertainty, including decisions on prioritisation and equitability, to help maximise the uptake of imminently available vaccines.

## Data and Code availability

The code for data analysis and figures generation are openly available at: https://gitlab.com/covid19aidashboard/covid-vaccination/ for reproducibility and transparent analysis. Our Twitter dataset was obtained using the publicly available Twitter API. Our Facebook dataset was obtained using the CrowdTangle platform, for which access must be requested from the organisation. All analysis was carried out in Python. Our code can be used as a reference for conducting similar studies. Due to the computationally expensive nature of the code, we recommend using a high performance computing resource.

## Data Availability

The code for data analysis and figures generation are openly available for reproducibility and transparent analysis. Our Twitter dataset was obtained using the publicly available Twitter API. Our Facebook dataset was obtained using the CrowdTangle platform, for which access must be requested from the organisation. All analysis was carried out in Python. Our code can be used as a reference for conducting similar studies. Due to the computationally expensive nature of the code, we recommend using a high performance computing resource.

https://gitlab.com/covid19aidashboard/covid-vaccination/

## Acknowledgements

The study reported here is funded by the Scottish Government Chief Scientist Office under its COVID-19 priority research programme (grant ref. COV/NAP/20/07) and is jointly led by co-Principal Investigators: AH and AS. AS is supported by BREATHE - The Health Data Research (HDR) Hub for Respiratory Health [MC_PC_19004], which is funded through the UK Research and Innovation Industrial Strategy Challenge Fund and delivered through HDR UK. AH is supported by the UK Government’s Engineering and Physical Sciences Research Council (EPSRC) grants (Ref. EP/T021063/1, EP/T024917/1).

## Abbreviations

BERT: Bidirectional Encoder Representations from Transformers
COVID-19: coronavirus disease
DL: Deep Learning
ML: Machine Learning
NLP: Natural Language Processing
P: p-value
VADER: Valence Aware Dictionary and sEntiment Reasoner

