## Appendix for "Artificial intelligence-enabled analysis of UK and US public attitudes on Facebook and Twitter towards COVID-19 vaccinations"

**Section-A1: Two-step thematic filtering strategy**

The first step filtering with COVID-19 related keywords utilised widely used terms from [19]: COVD19, CoronavirusPandemic, COVID-19, 2019nCoV, CoronaOutbreak, coronavirus, WuhanVirus, covid19, coronavirus pandemic, covid-19, 2019ncov, corona outbreak and wuhanviru.  The vaccination terms used for second step thematic filtering were selected by our team: vaccine, vaccination, immunise, immunize, immunisation and immunization. The two-step thematic filtering process was applied using the above mentioned keywords before processing and analysis.

**Section-A2: Additional Analysis Techniques:**

- **Averaged time series sentiment trends**: The time series sentiment represents the temporal distribution of averaged weekly sentiment in the UK and US. This was analysed over the period of study (March-Nov 2020).
- **Sentiment word clouds and text N-gram analysis**: Sentiment word clouds are graphical representations of the most frequently occurring words and multi-word expressions, and their associated sentence-level sentiments. The word font size is proportional to the frequency of its occurrence, within each positive, negative and neutral word cloud. Sentiment word clouds were generated and analysed for the positive, negative and neutral sentiments, based on the top frequent N-grams (unigrams, bigrams and trigrams).
- **Geo-spatial sentiment analysis:** The geo-spatial distribution of averaged UK and US sentiment was mapped and analysed.
- **Statistical analysis**: Pearson r correlation values were conducted between sentiments and date, to demonstrate the trends of sentiments across time statistically.
- **Independent Surveys**: Related public surveys from the UK and US over the period of study, were analysed to compare and contrast findings with our social-media analysis.

Sentiment word cloud and N-gram analysis was applied to specific time-periods of interest, around points of inflexion on sentiment trend graphs, to identify topics of discussion and glean insight into the positive and negative content of online discourses. The analysis was also carried out over the full period of study, March to November 2020, to identify underlying themes and topics. Findings were validated, and further insights obtained, through a manual reading by our team, of randomly selected social-media posts around target points of interest. Relevant social-media datasets and outputs were anonymised, and statistical aggregates presented through a publicly available dashboard [24].

**Section-A3: Statistical Analysis of Sentiment Trends**

Pearson's r for positive, negative and neutral public sentiments on Twitter, for the UK, was approximately zero for each case, and the *P* value was not significant. On the other hand, positive, negative and neutral sentiment trends on Twitter for the US had r values of 0.77 (*P* < 0.001), -0.62 (*P* < 0.001) and -0.6 (*P* < 0.001) respectively, which represent an increase in positive sentiment and decrease in both negative and neutral sentiments over time.

Pearson’s r for positive UK public sentiment on Facebook, had a value of -0.62 (*P* < 0.001) which represents a decrease in positive sentiment trend. Negative UK sentiments on Facebook had an r value of 0.26 with a p-value that was not significant, whilst neutral UK sentiments had an r value of 0.66 (*P* < 0.001), representing an increase in neutral sentiment over time. Similarly, positive sentiment trend on Facebook for the US, had an r value of -0.56 (*P* < 0.001), signifying a decrease in positive sentiment over time. Pearson’s r for negative US sentiment on Facebook was 0.15 with a p-value that was not significant, whereas neutral US sentiment had an r value of 0.56 (*P* < 0.001).

**Section-A4: Sentiment Word Clouds and Text N-Gram Analysis**

We identified and analysed notable events that were of interest to social media users, and summarise them below (some of these were also identified in the analysis on the sentiment trend graphs in Figures 2 and 3).

- First human trial vaccination: Towards the start of the pandemic (23 April), there were a number of social-media conversations around the first human vaccine trials. Many users indicated a positive attitude towards them.
- UK government rejects EU COVID-19 vaccination scheme: User posts indicated negative sentiment towards the UK government's decision to opt out of the EU vaccination scheme (10 July).
- Russian vaccine:  Tweets relating to the deployment of a Russian vaccine had been increasingly prominent, since their government approved a vaccine without large scale testing (11 August). Public sentiment towards this was divisive.
- Oxford Vaccine trial paused: Following the Oxford Vaccination trial being paused due to reports surrounding side effects (9 September). Sentiments towards this were predominantly negative.
- President Trump vaccine statement: President Trump claims that a vaccine could be ready within 3-4 weeks (16 September), and also referred to herd immunity as the “herd mentality”, which proved mockery on Twitter.
- Pfizer announcement: Pfizer and BioNTech announce 90% effectiveness of their vaccine, in preventing COVID-19 (9 November). Public sentiment has been markedly positive in both the UK and the US, on Twitter, whilst on Facebook there has been a marked increase in neutral sentiment since the announcement.

Visual inspection of the sentiment word clouds for the US (Figures A2 and A3 in Appendix) indicate that the public reacted negatively to the news of "Trump" trying to buy exclusive rights to the COVID-19 vaccine.  An “anti-Russian” sentiment was also identified, arising from concerns raised by the National Institutes for Health, along with a number of other healthcare organisations, on the development of the Russian vaccine (on 11 August 2020), and from claims that fundamental parts of the safety process were missed during its development.

Overall, the word clouds indicated positive public sentiments towards promising trials for the COVID-19 vaccine. Similarly, there is a negative sentiment related to the pausing of Oxford trials with a concern for vaccine safety issues. In general, however, public sentiments on vaccine trials e.g. trials at Oxford and their resumption is positive.

The sentiment word clouds for the UK tweets (Figure A2) prominently featured words including: UK, world, trial, new and people. Oxford features frequently in tweets with a positive sentiment. Tweets with a negative sentiment frequently used words including: people, flu, get, UK and US.  Words such as UK, Oxford and trial also feature prominently in neutral sentiment tweets. For tweets in the US (Figure A.3), words that were frequent for both positive and negative sentiment word clouds include: people and US. "Trial" is frequent in both positive and neutral sentiment word clouds, which is similar to the UK. The word Trump appears frequently in tweets with negative sentiments. Interestingly, India's Serum Institute appeared frequently in neutral sentiment tweets. Overall, there was some similarity in the words that appeared in the UK and the US. However, the frequency of the word "trial" when used positively was greater in the UK than in the US. "Oxford" was not used frequently in tweets from the US, indicating that, as can be expected, public discussion surrounding the Oxford vaccination trials is primarily centred in the UK.

An analysis of the most frequent positive, negative and neutral ‘*bigrams*’ used across Facebook and Twitter (in both the UK and US) was carried out in relation to COVID-19 vaccination (Table A1 in Appendix). The most frequent positive bigram was “public health”, and the most frequent negative bigram was “flu vaccine”. Bill Gates appeared in a plethora of conspiracy theories associated with the COVID-19 vaccine. The conspiracies, which largely related to corporate profiteering by making a vaccine mandatory or to instituting corporate control by installing microchips through the vaccination process, generated the frequent negative bigram “Bill Gates”, as well as the word “Gates” in word clouds associated with a negative public sentiment. Although many of the conspiracies have been debunked, they nevertheless represent a window into public psyche and mass fears and concerns related to the COVID-19 vaccine.

The most frequent positive ‘*trigrams*’ (Table A2 in Appendix) included “disease control prevention”, “Dr. Anthony Fauci” and “World Health Organisation”. Note, Dr Anthony Fauci is an American physician and immunologist, who had predicted that a vaccine might be widely available in the US by April 2021. The prominence of the “World Health Organisation” in the positive category is promising, and it would be interesting to further explore public trust towards it. In contrast, the most frequent negative words included “centers disease control”, “President Donald Trump” and “President Rodrigo Duterte”, the latter two indicating the importance of political figures in public attitude.

**Section-A5:** **Overall Averaged Sentiment Analysis**

The averaged positive sentiment in the UK was 54.2% on Twitter and 61.1% on Facebook, giving an overall averaged positive sentiment of 57.7% across the two platforms. Similarly, the averaged positive sentiment in the US was found to be comparable: 51.5% on Twitter and 62.1% on Facebook, with an overall averaged positive sentiment of 56.8%.

We found that Twitter users expressed a relatively higher, averaged negative sentiment and a lower positive sentiment compared to Facebook users, for both the UK and the US. In the UK, the overall averaged negative sentiment related to COVID-19 vaccination was 27.95% on Twitter and 17.04% on Facebook. Similarly, in the US, the overall averaged negative sentiment was 30.57% on Twitter and 17.73% on Facebook. This appeared to be consistent with the study in [17], which found public opinion on Twitter to be often more negatively biased than on Facebook, where it is more positively biased.

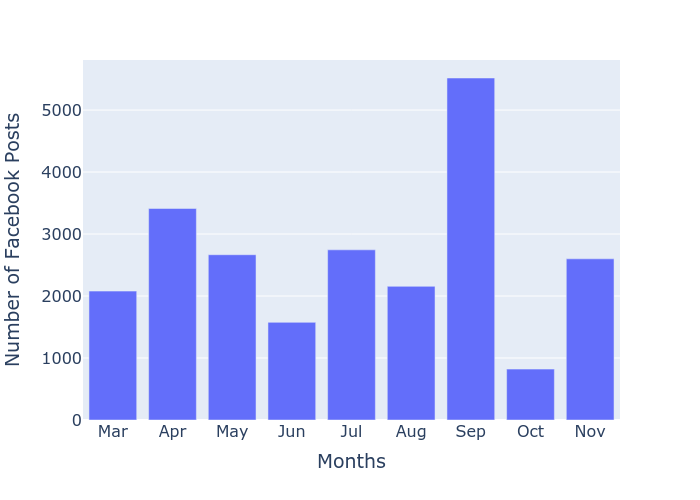

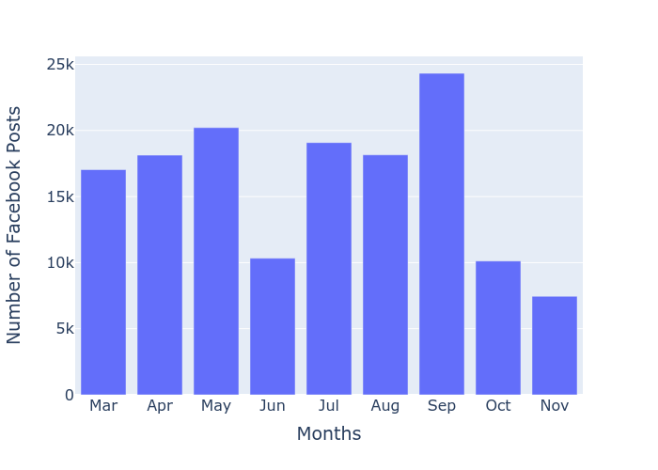

**a.(Facebook UK)	b.(Facebook US)**

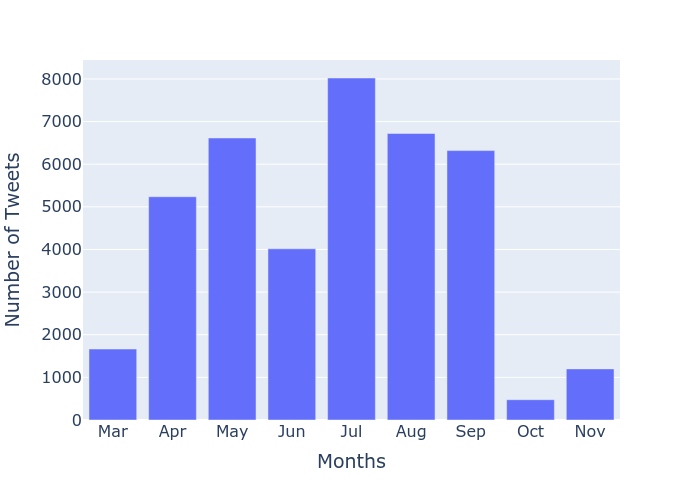

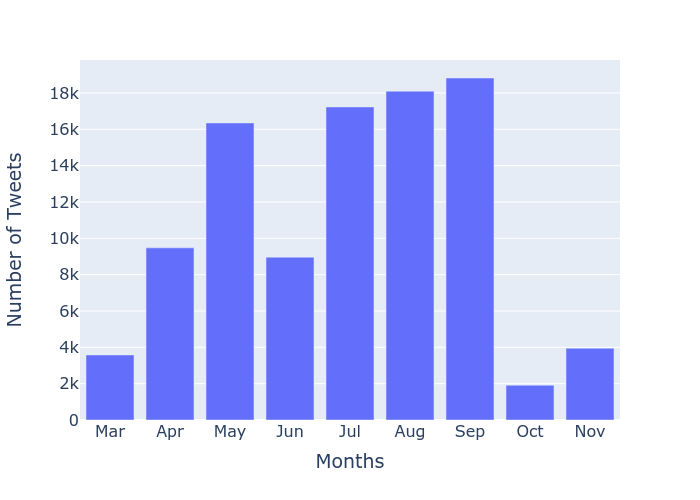

**c.(Twitter UK)	d.(Twitter US)**

**Figure-A1: Monthly Volume Trends - total number of social-media posts identified after two-step filtering, by platform (Facebook (a)(b), Twitter (c)(d)) and, country (UK, US)**

**UK (i)**

**Positive	Neutral	Negative**

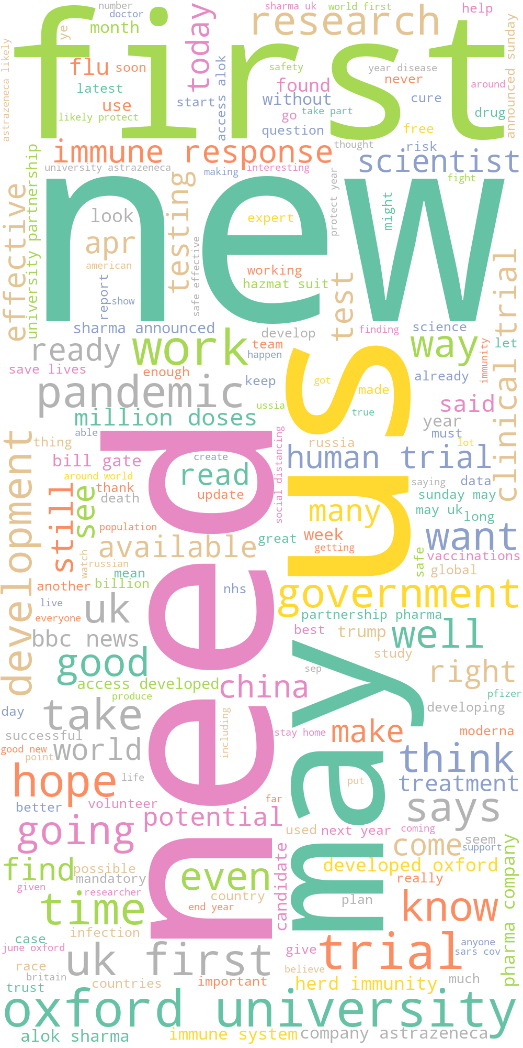

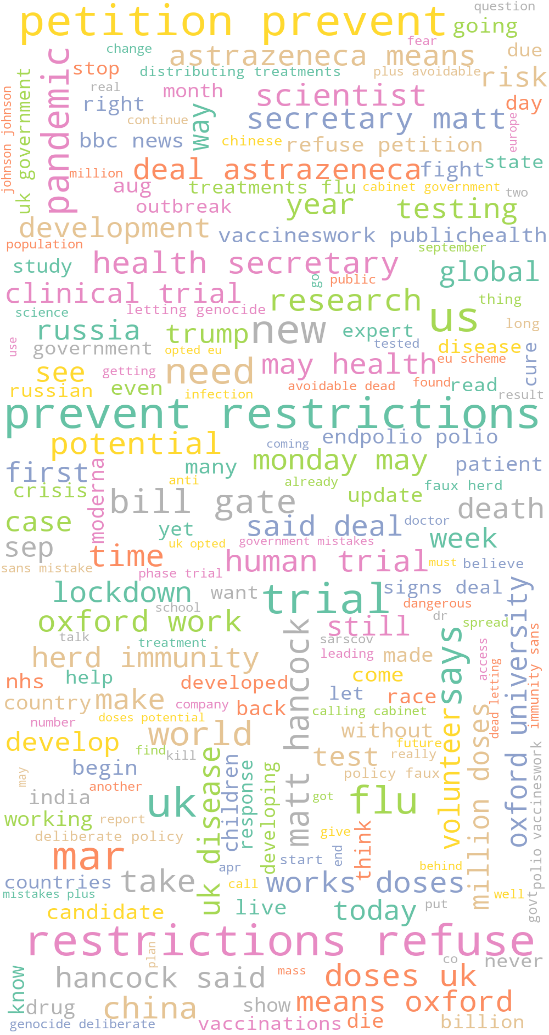

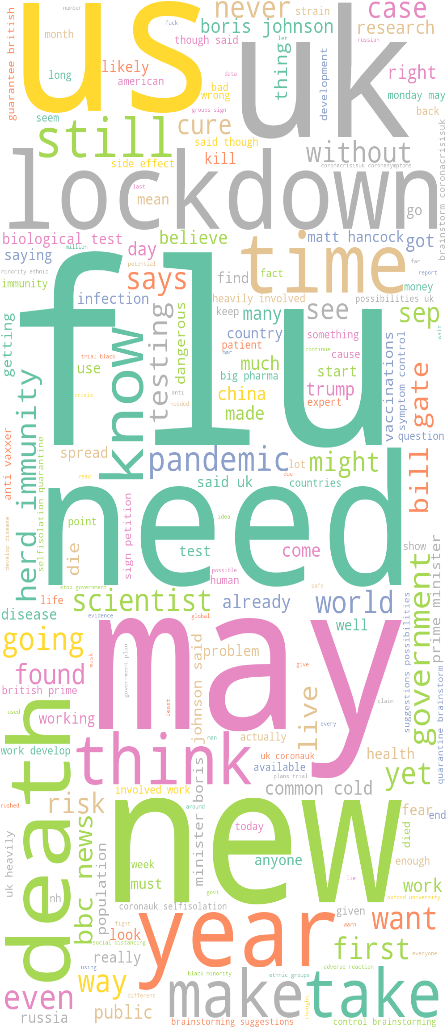

**US (ii)**

**Positive	Neutral	Negative**

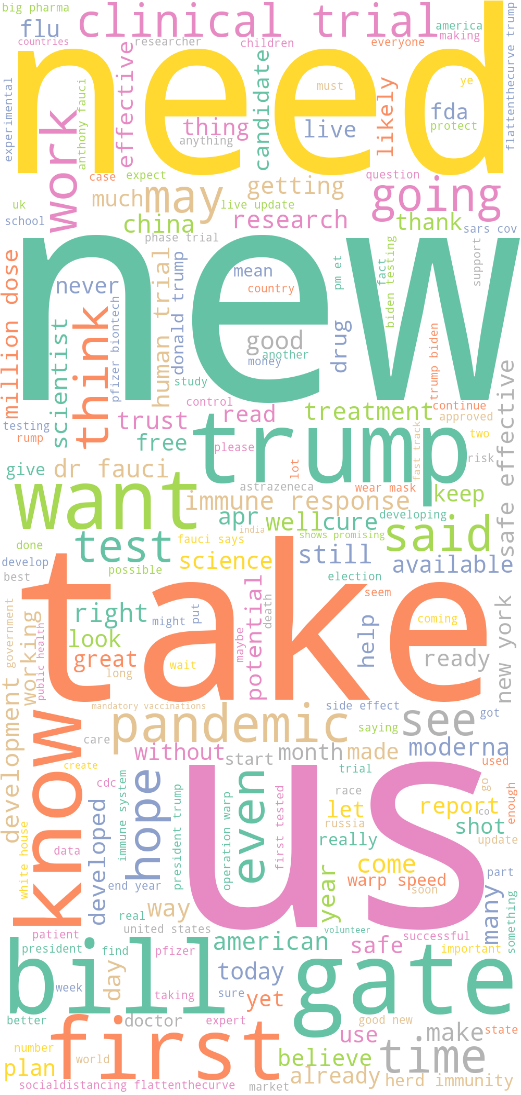

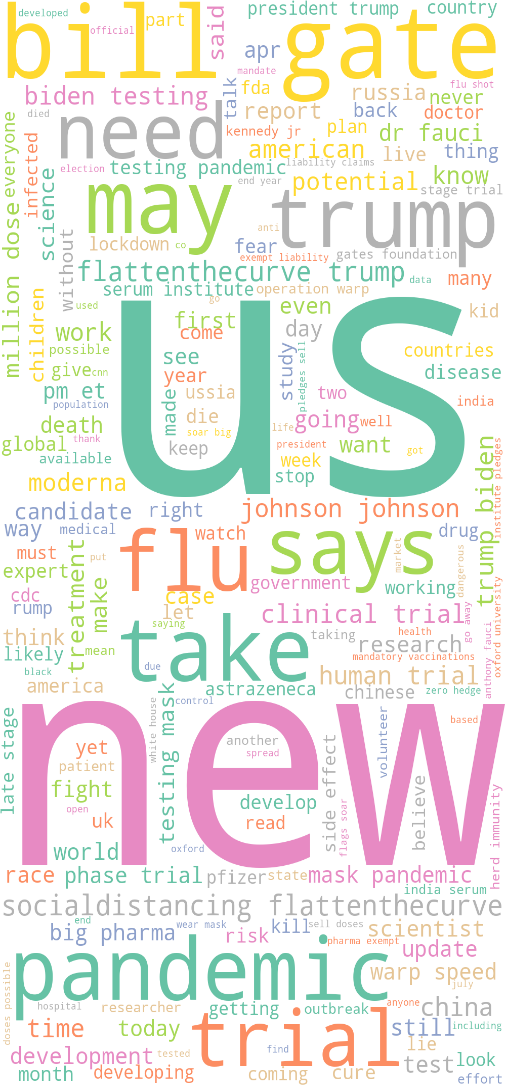

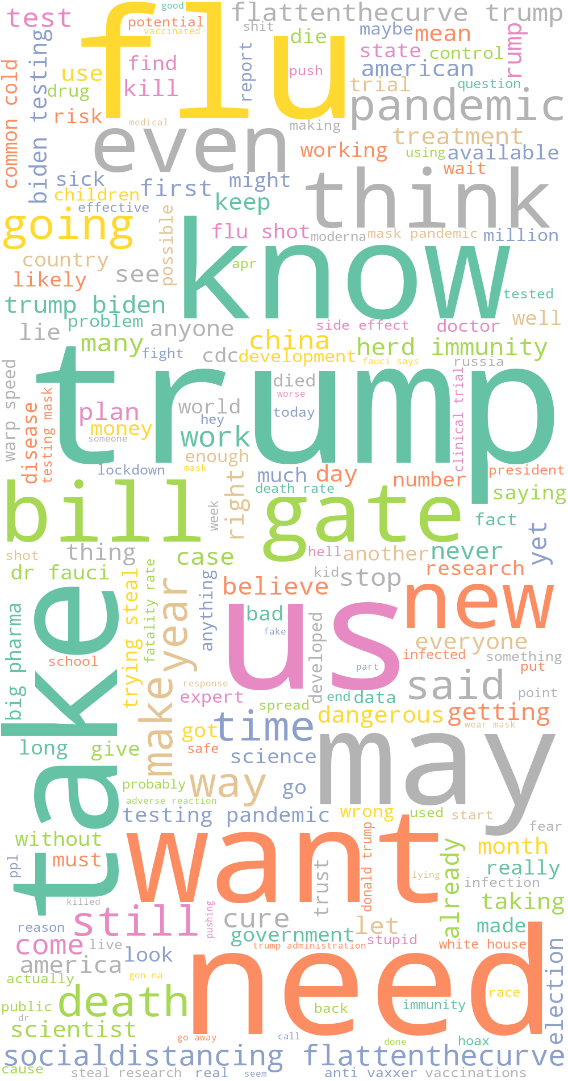

**Figure-A2: Twitter (Overall) Word Cloud: (i) UK (ii) US**

**UK (i)**

**Positive	Neutral	Negative**

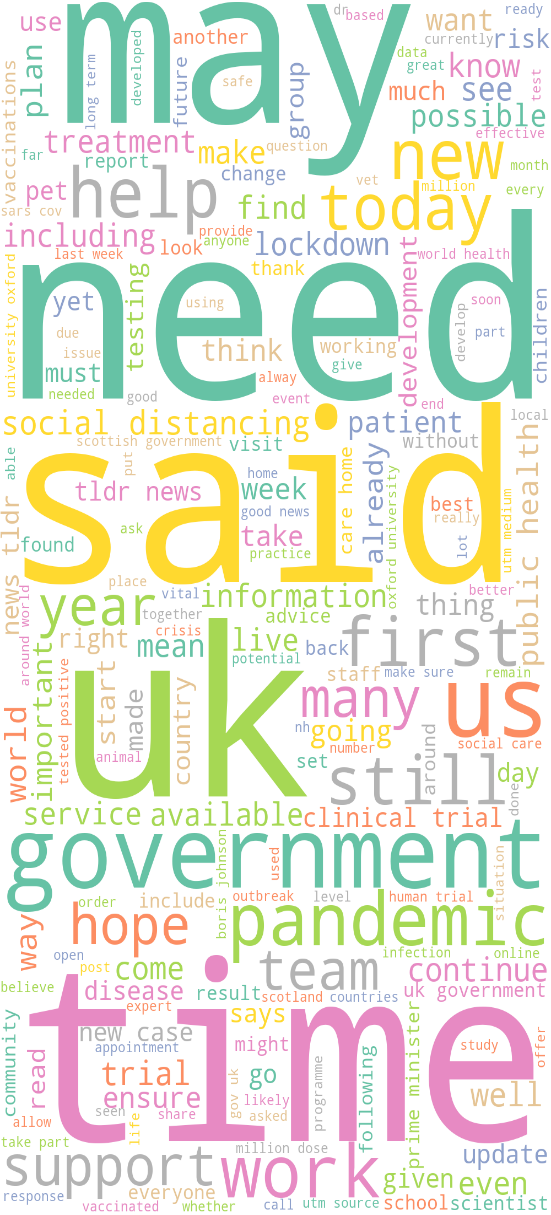

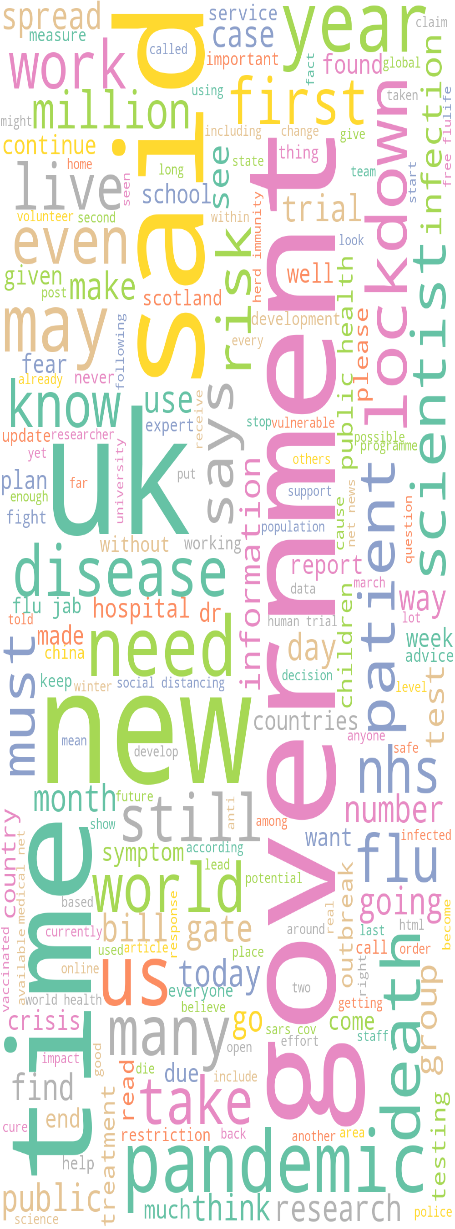

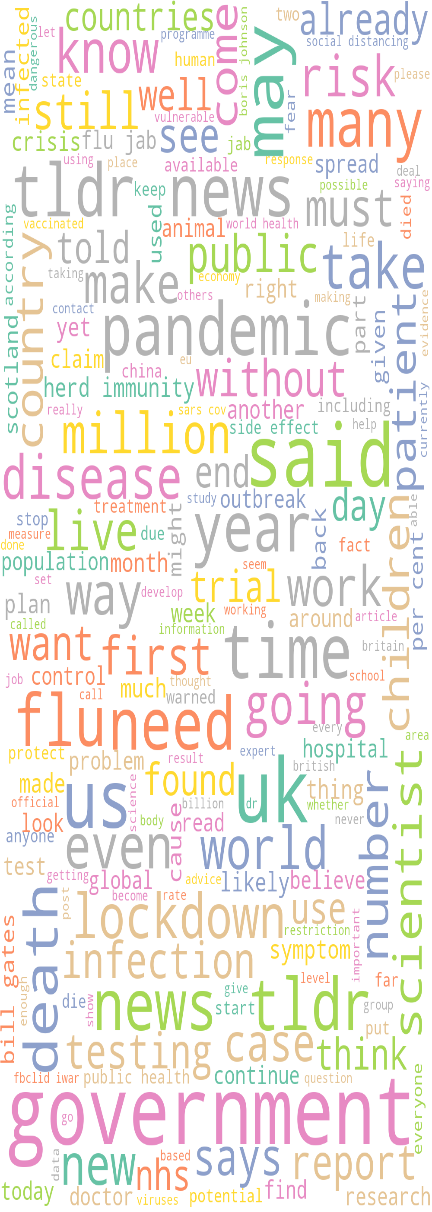

**US (ii)**

**Positive	Neutral	Negative**

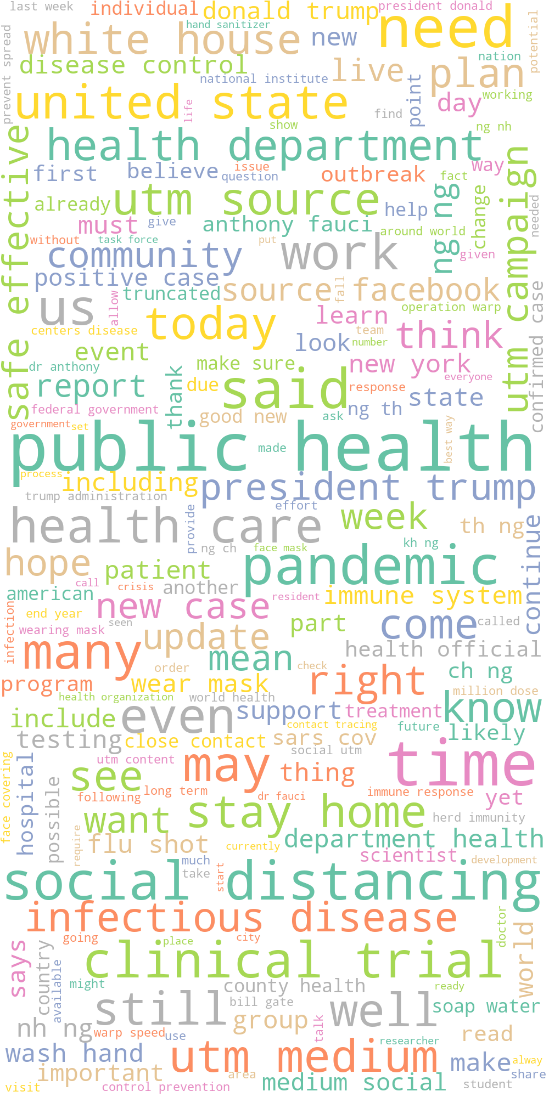

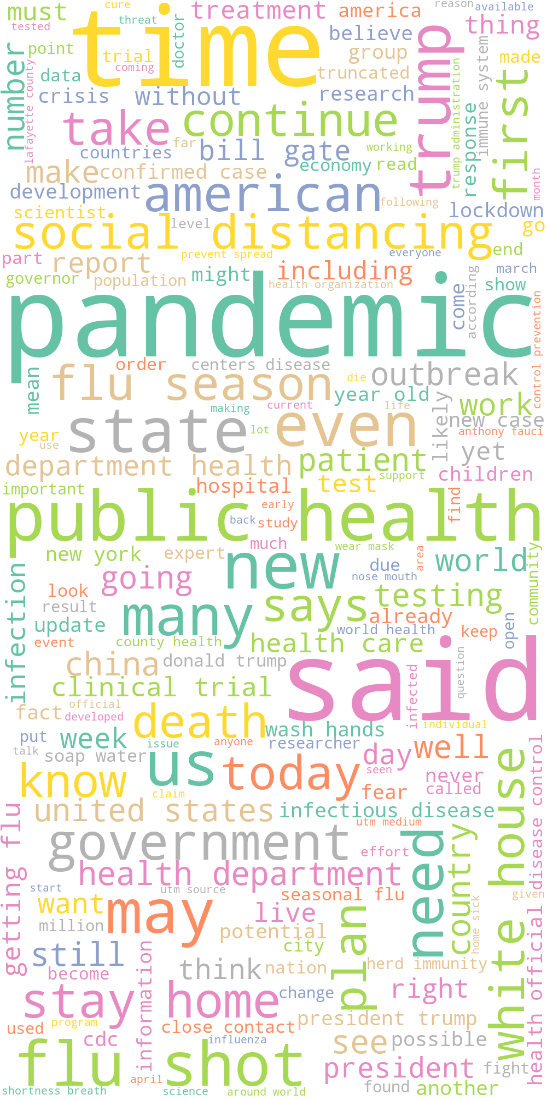

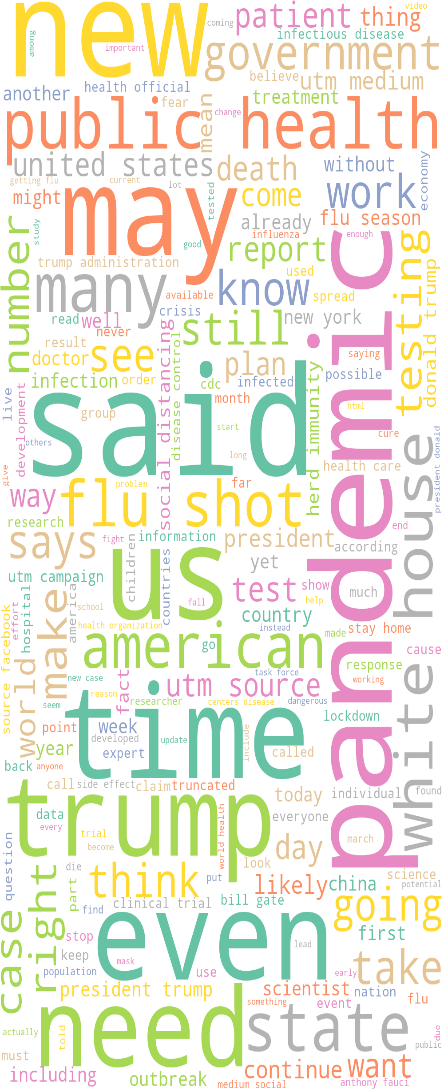

**Figure-A3: Facebook (Overall) Word Cloud: (i) UK (ii) US**

**Table-A1: Most Frequent Bi-grams (across Facebook and Twitter in the UK and US)**

| **Positive Sentiment** | **Neutral Sentiment** | **Negative Sentiment** |
| --- | --- | --- |
| Public health | refuse covid | flu shot |
| social distance | restrictions refuse | covid vaccines |
| healthcare | prevent restrictions | covid coronavirus |
| covid pandemic | petition prevent | herd immunity |
| stay home | oxford vaccine | vaccine trial |
| white house | stay home | herd immunity |
| clinical trials | social distancing | president trump |
| safe effect | covid pandemic | get flu |
| Bill gates | Flu season | Social distancing |
| Health department | Get flu | Vaccine trials |
| vaccine available | matt hancock | flu season |

**Table-A2: Most Frequent Tri-grams (across Facebook and Twitter in the UK and US)**

| **Positive Sentiment** | **Neutral Sentiment** | **Negative Sentiment** |
| --- | --- | --- |
| dr anthony fauci | get flu shot | centers disease control |
| operation warp speed | centers disease control | world health organization |
| centers disease control | world health organization | president donald trump |
| medium social utm | disease control prevention | covid vaccine trials |
| disease control prevention | get flu vaccine | get covid vaccine |
| county health department | dr anthony fauci | disease control prevention |
| covid vaccine trial | getting flu vaccine | dr anthony fauci |
| world health organization | county health department | medium social utm |
| health human services | stay home sick | trump biden testing |
| food drug administration | getting flu shot | operation warp speed |
